# Variations in Personal Protective Equipment Preparedness in Intensive Care Units during the COVID-19 Pandemic: A Survey of Asia-Pacific Countries: On Behalf of the SPARTAN Collaborative - Small Projects, Audits, Research Trials - Australia/New Zealand

**DOI:** 10.1101/2020.05.06.20093724

**Authors:** Arvind Rajamani, Ashwin Subramaniam, Kiran Shekar, Jumana Haji, Jinghang Luo, Shailesh Bihari, Wai Tat Wong, Navya Gullapalli, Markus Renner, Claudia Maria Alcancia, Kollengode Ramanathan

## Abstract

**Objectives:** To evaluate PPE-preparedness across intensive care units (ICUs) in 6 Asia-Pacific countries. PPE-preparedness was defined as the adherence to guidelines, training HCWs, procuring PPE stocks and responding appropriately to a suspected case (transportation and admission to hospital).

**Design:** Cross-sectional web-based survey.

**Setting:** ICUs in Australia, New Zealand (NZ), Singapore, Hong Kong (HK), India and Philippines with a 24/7 Emergency/Casualty Department, and capable of mechanically ventilating patients for >24 hours.

**Interventions:** Questionnaire sent to intensivists in 633 Level ll/lll ICUs in 6 Asia-Pacific countries by email, WhatsApp™ and text messaging.

**Main outcome measures:** 263 intensivists responded, of whom 231 were eligible for analysis. Response rates were 68%-100% in all countries except India, where it was 24%. 97% either conformed to or exceeded WHO recommendations for PPE-practice. 59% employed airborne precautions irrespective of aerosol-generation-procedures. There were variations in negative-pressure room use (highest in HK/Singapore), training (best in NZ), and PPE stock-awareness (best in HK/Singapore/NZ). High-flow-nasal-oxygenation and non-invasive ventilation were not options in most HK (66.7%, 83.3% respectively) and Singapore ICUs (50%, 80% respectively), but were considered in other countries to a greater extent. 38% reported not having specialized airway teams. Showering and “buddy-systems” were underutilized. Clinical waste disposal training was suboptimal (38%).

**Conclusions:** Most intensivists from six Asia-Pacific countries appeared to be aware of the WHO PPE-guidelines by either conforming to/exceeding the recommendations. Despite this, there were widespread variabilities across ICUs and countries in several domains, particularly related to PPE-training and preparedness. Standardising PPE guidelines may translate to better training, better compliance and policies that improve HCW safety. Adopting low-cost approaches such as buddy-systems should be encouraged. More importantly, better pandemic preparedness and building systems with deeply embedded culture of safety is essential to ensure the safety and well-being of HCWs during such pandemics.

**Author Contributorship:** 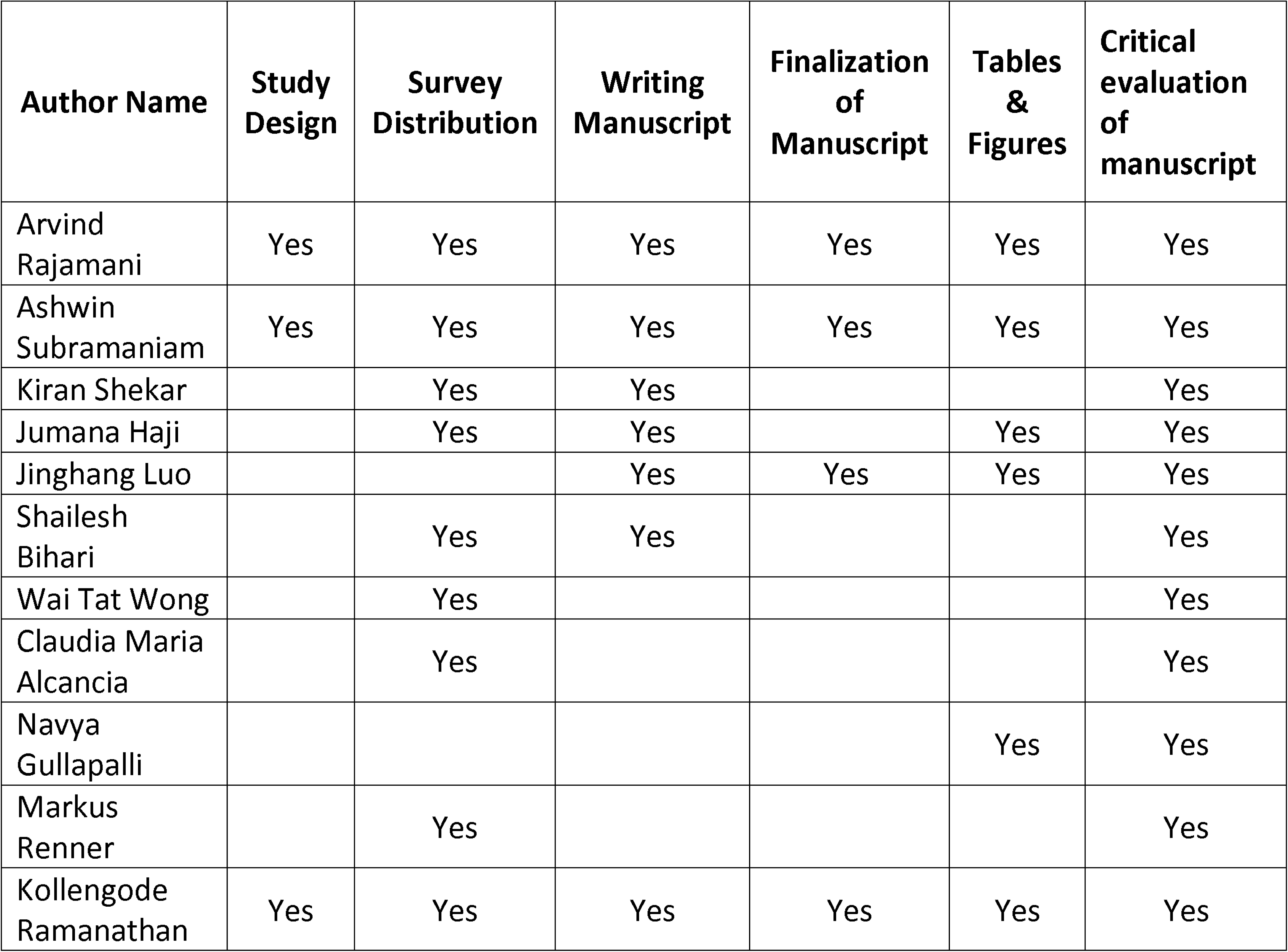

**Summary Box:** *What is already known on this topic:*

- Personal-protective equipment (PPE) is the cornerstone to preventing HCW- infections. A search was done on March 23, 2020 on PubMed, Embase or Google Scholar using the mesh terms “personal protective equipment”, “PPE”, “preparedness OR practice OR training”. It revealed no previous studies on PPE preparedness in intensive care units (ICUs). No filters were used for the search.
- Several guidelines/recommendations issued by health organisations on PPE practice exist

*What are the new findings:*

- As the first study to evaluate PPE-preparedness in ICUs, it demonstrated major concerns on PPE-preparedness across several ICUs, particularly in Australia, India and Philippines. There was suboptimal PPE-training, under-utilisation of low-cost interventions such as buddy-systems/team-training, and stock-awareness.
- The guidelines by health organisations on PPE practice have several conflicting recommendations.

*How might it impact on clinical practice in the foreseeable future:*

- Standardising PPE guidelines by health organisations may translate to better training, better compliance and policies that improve HCW safety.
- To ensure the safety and well-being of HCWs, urgent measures are needed to improve PPE-preparedness and building systems with deeply embedded culture of safety. By helping ICUs evaluate and improve their current state of PPE preparedness, the study may help prevent healthcare worker infections and save lives.

## Main Manuscript

## Introduction

The Severe Acute Respiratory Syndrome-Coronavirus-2 (SARS-CoV-2) pandemic has caused an unprecedented rise in the number of hospital and intensive care unit (ICU) admissions worldwide. Between 5-32% of coronavirus disease-2019 (COVID-19) patients may require critical care support.[1,2] Asia-Pacific countries are preparing for an anticipated surge in ICU admissions.[1,3,4] Due to its high transmissibility,[5] ICU healthcare workers (HCWs) are at particular risk of infection.[6]

Adequate preparedness with personal-protective equipment (PPE) is the cornerstone to preventing HCW-infections.[7,8] PPE-preparedness for pandemics was defined as the adherence to guidelines, training HCWs, procuring PPE stocks and responding appropriately to a suspected case (transportation and admission to hospital).[9] Suboptimal PPE-preparedness may cause PPE breaches, thereby exposing HCWs to SARS-CoV-2 infection.[10] Reports in India and Australia suggest a rising rate of HCW infections despite a low overall caseload,[11,12] raising concerns about the effectiveness of PPE practice and stocks.[13,14] Moreover, there are conflicting recommendations from international, national, and regional organisations.[5,15] For example, the World Health Organisation (WHO) recommends a tiered approach of using droplet precautions for non-aerosol-generating procedures (AGPs), and using airborne precautions only for AGPs. However, the Australia-New Zealand Intensive Care Society (ANZICS)[15] recommends that ICU-HCWs must routinely use airborne precautions, irrespective of the AGP-risk. HCW-infection rates vary between countries,[16,17] with reports suggesting a lower incidence with advanced PPE measures and good PPE-practice bundles.[18-20]

While HCWs deserve the highest level of PPE-protection, inappropriate PPE usage driven by anxiety rather than clinical requirements, may deplete already scarce stocks. PPE availability must be accompanied by rigorous training systems in every ICU.[21-27] Since there is no literature on PPE-preparedness for the COVID-19 pandemic, we conducted a multinational cross-sectional survey of intensivists in 6 Asia-Pacific countries to comprehensively evaluate PPE-preparedness and compliance with WHO PPE-recommendations.

## Methods

### Study Design

Cross-sectional web-based survey of intensivists to evaluate PPE-preparedness in Asia-Pacific ICUs. The survey remained open for completion between 25/03/2020 and 06/04/2020.

### Development of the survey

Established standards and guidelines were utilized to design and validate the survey prior to dissemination.[28,29] The broad research topic (PPE-preparedness in Asia-Pacific ICUs) and the questionnaire were developed after several rounds of consensus-building process between intensive care and infectious diseases specialists, based on data from the EuroNHID project.[7] Since it was a multinational survey, the WHO recommendations^9^ were chosen as the reference standard to evaluate PPE-preparedness in each country. After a process of item-generation and item-reduction, the questionnaire was piloted using 6 intensivists to evaluate completeness and clarity, and then administered to 12 intensivists from participating countries to assess its ability to discriminate among responses, ease of use and content validity (Table 1).

**Table 1.**
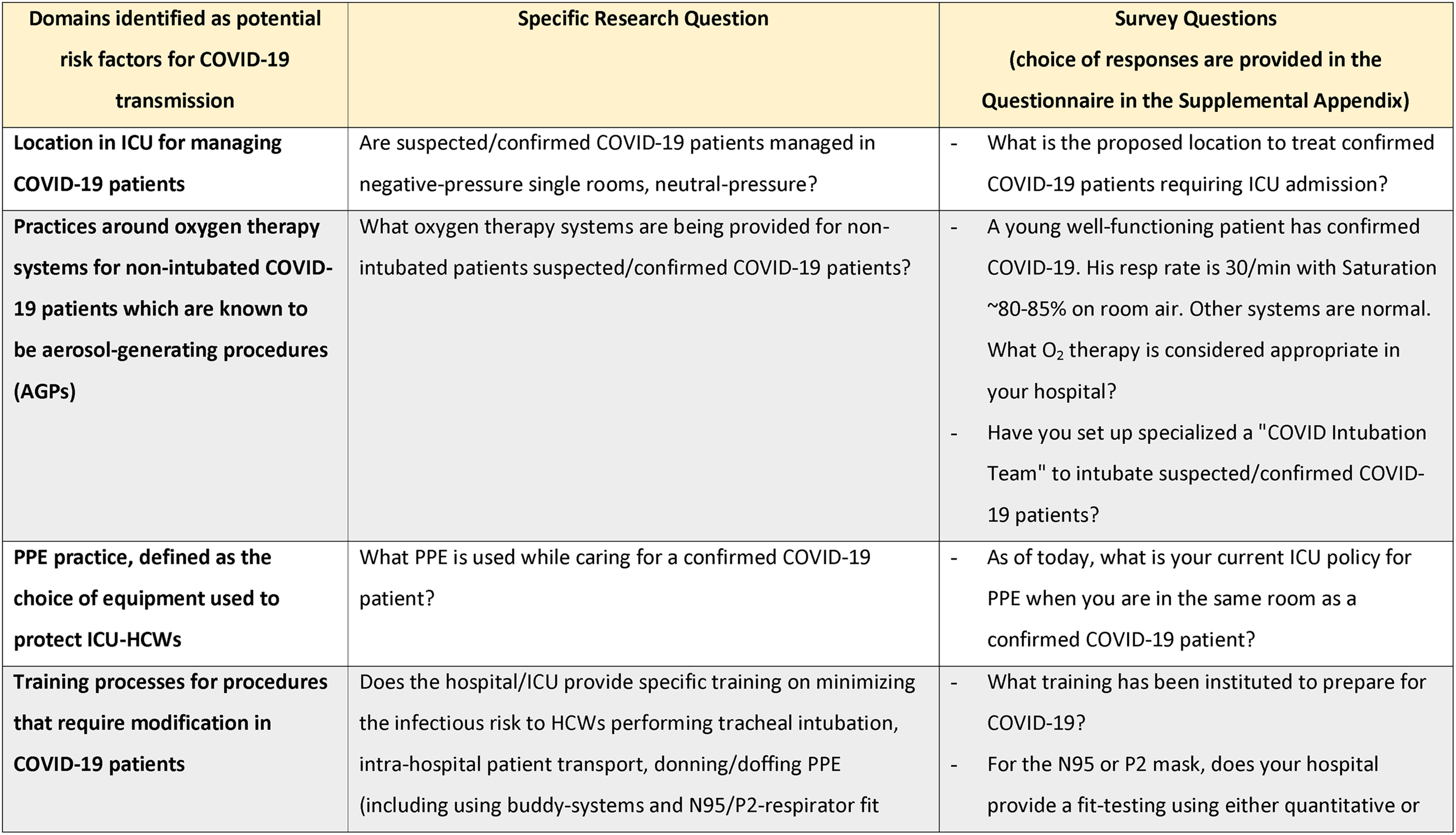
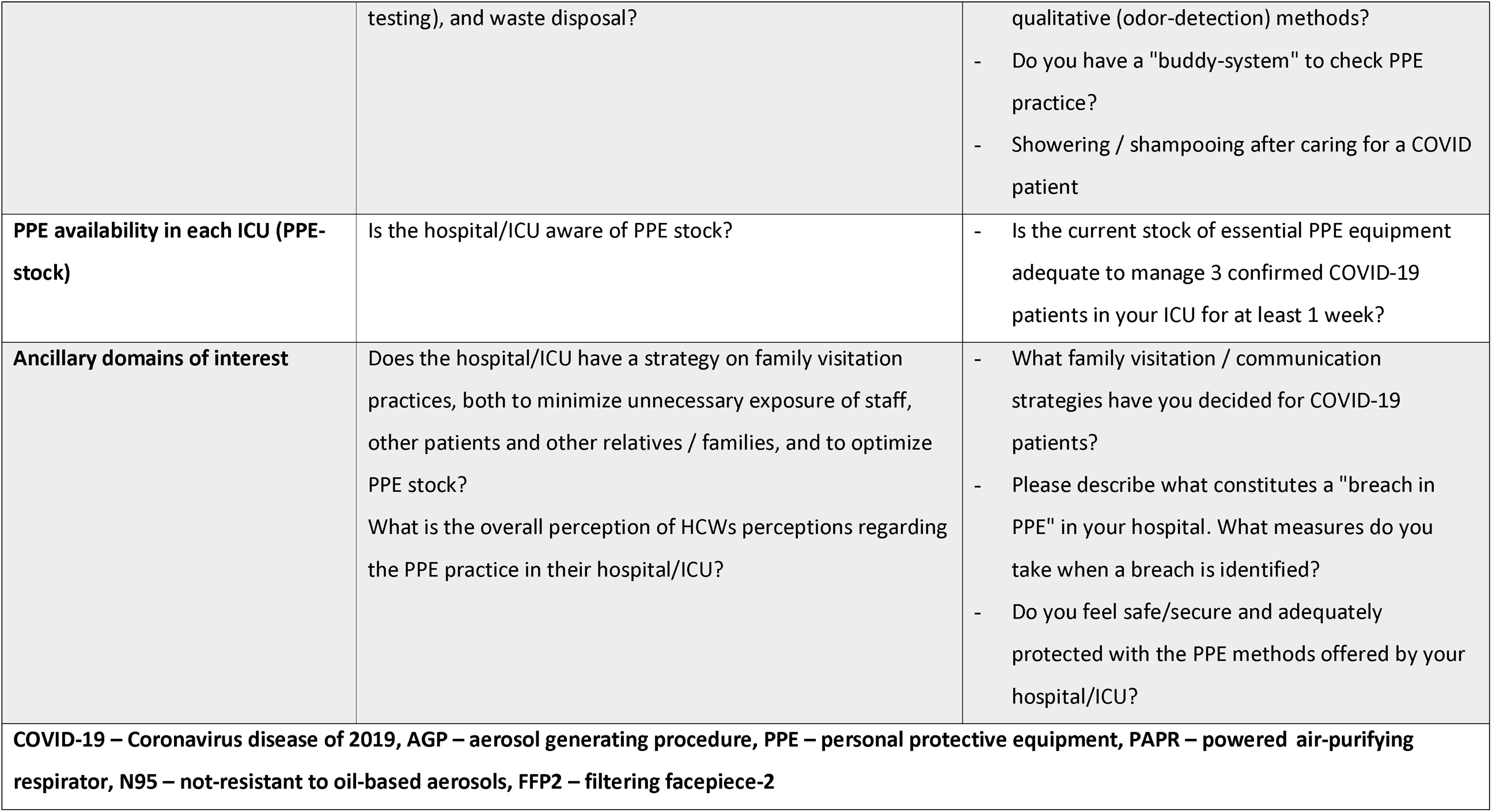
Design and Development of the Questionnaire

| **Domains identified as potential risk factors for COVID-19 transmission** | **Specific Research Question** | **Survey Questions**<br>**(choice of responses are provided in the Questionnaire in the Supplemental Appendix)** |
| --- | --- | --- |
| **Location in ICU for managing COVID-19 patients** | Are suspected/confirmed COVID-19 patients managed in negative-pressure single rooms, neutral-pressure? | - What is the proposed location to treat confirmed COVID-19 patients requiring ICU admission? |
| **Practices around oxygen therapy systems for non-intubated COVID-19 patients which are known to be aerosol-generating procedures (AGPs)** | What oxygen therapy systems are being provided for non-intubated patients suspected/confirmed COVID-19 patients? | - A young well-functioning patient has confirmed COVID-19. His resp rate is 30/min with Saturation ~80-85% on room air. Other systems are normal. What O<sub>2</sub> therapy is considered appropriate in your hospital?<br>- Have you set up specialized a "COVID Intubation Team" to intubate suspected/confirmed COVID-19 patients? |
| **PPE practice, defined as the choice of equipment used to protect ICU-HCWs** | What PPE is used while caring for a confirmed COVID-19 patient? | - As of today, what is your current ICU policy for PPE when you are in the same room as a confirmed COVID-19 patient? |
| **Training processes for procedures that require modification in COVID-19 patients** | Does the hospital/ICU provide specific training on minimizing the infectious risk to HCWs performing tracheal intubation, intra-hospital patient transport, donning/doffing PPE (including using buddy-systems and N95/P2-respirator fit | - What training has been instituted to prepare for COVID-19?<br>- For the N95 or P2 mask, does your hospital provide a fit-testing using either quantitative or |
|  | testing), and waste disposal? | qualitative (odor-detection) methods?<br>- Do you have a "buddy-system" to check PPE practice?<br>- Showering / shampooing after caring for a COVID patient |
| **PPE availability in each ICU (PPE-stock)** | Is the hospital/ICU aware of PPE stock? | - Is the current stock of essential PPE equipment adequate to manage 3 confirmed COVID-19 patients in your ICU for at least 1 week? |
| **Ancillary domains of interest** | Does the hospital/ICU have a strategy on family visitation practices, both to minimize unnecessary exposure of staff, other patients and other relatives / families, and to optimize PPE stock?<br>What is the overall perception of HCWs perceptions regarding the PPE practice in their hospital/ICU? | - What family visitation / communication strategies have you decided for COVID-19 patients?<br>- Please describe what constitutes a "breach in PPE" in your hospital. What measures do you take when a breach is identified?<br>- Do you feel safe/secure and adequately protected with the PPE methods offered by your hospital/ICU? |
| **COVID-19 – Coronavirus disease of 2019, AGP – aerosol generating procedure, PPE – personal protective equipment, PAPR – powered air-purifying respirator, N95 – not-resistant to oil-based aerosols, FFP2 – filtering facepiece-2** |  |  |

### Participant population and distribution of the survey

Following ethical approval (approval number 2020/ETH00705) from local Human Research Ethics Committee, the survey weblink was distributed by email, text message and WhatsApp™ to qualified intensivists in Australia, New Zealand (NZ), Singapore, Hong Kong (HK), India and Philippines. The target participants were intensivists from hospitals with a 24/7 Emergency/Casualty Department, having an ICU capable of supporting mechanically ventilated patients for >24 hours.[30,31] Two reminders were sent 3 days apart. One response per ICU was allowed. If multiple responses per site were received, the first response was chosen. Participation was voluntary, with no incentives offered, financial or otherwise.

## Results

The survey was administered to 633 intensivists from Australia (n=99), NZ (n=14), HK (n=13), Singapore (n=6), India (n=481) and Philippines (n=20). The response rate was 100% in NZ and Singapore, 92.3% in HK (12/13), 69% (68/99) in Australia, 80% in Philippines (16/20) and 24% in India (115/481). Overall, 263/633 intensivists responded (42%). After exclusion of duplicates and responses from ineligible institutions, 231 (37%) valid responses were included in the final analysis (CONSORT diagram, Figure 1). Except in Philippines, responses in each country had wide geographical spread across states/territories/regions.

**Figure 1.**
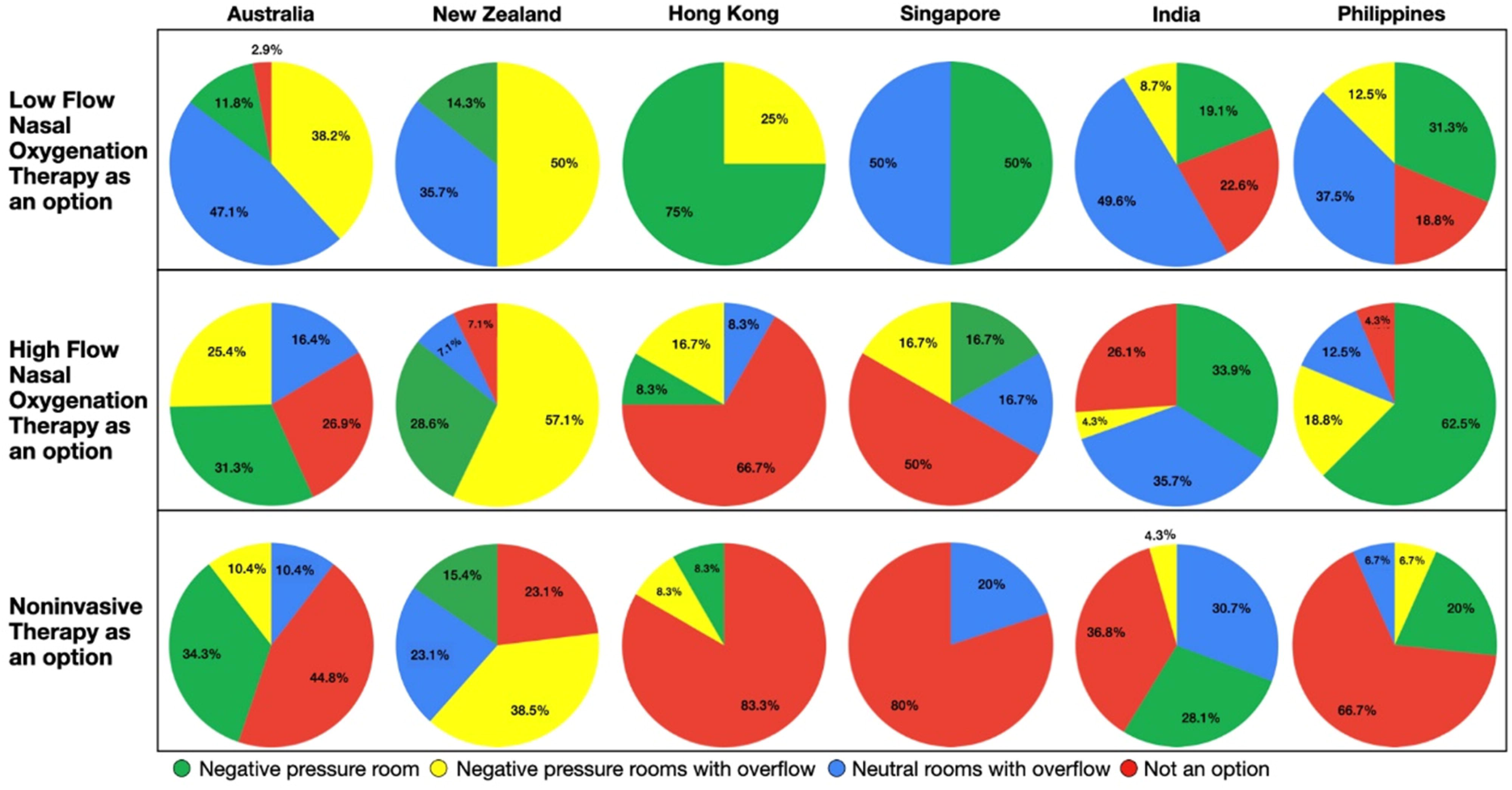
CONSORT diagram demonstrating 42% response rate. After exclusion, 231 ICUs were included for final analysis. Overall response rate was very good, except in India, which reduced the overall response rate. Key: ICU - intensive care unit

### PPE-training (Table 2 and Table 3)

Regular team-training on tracheal intubation was provided in 79% NZ ICUs, 59% in Australia, 50% in Singapore, and 19%-33% in the other countries. Overall, 66% reported establishing special intubation teams of senior anaesthetists (4%), senior intensivists (10%), or a combination of both (52%), ranging from 33% in Singapore to 93% in NZ.

**Table 2:**
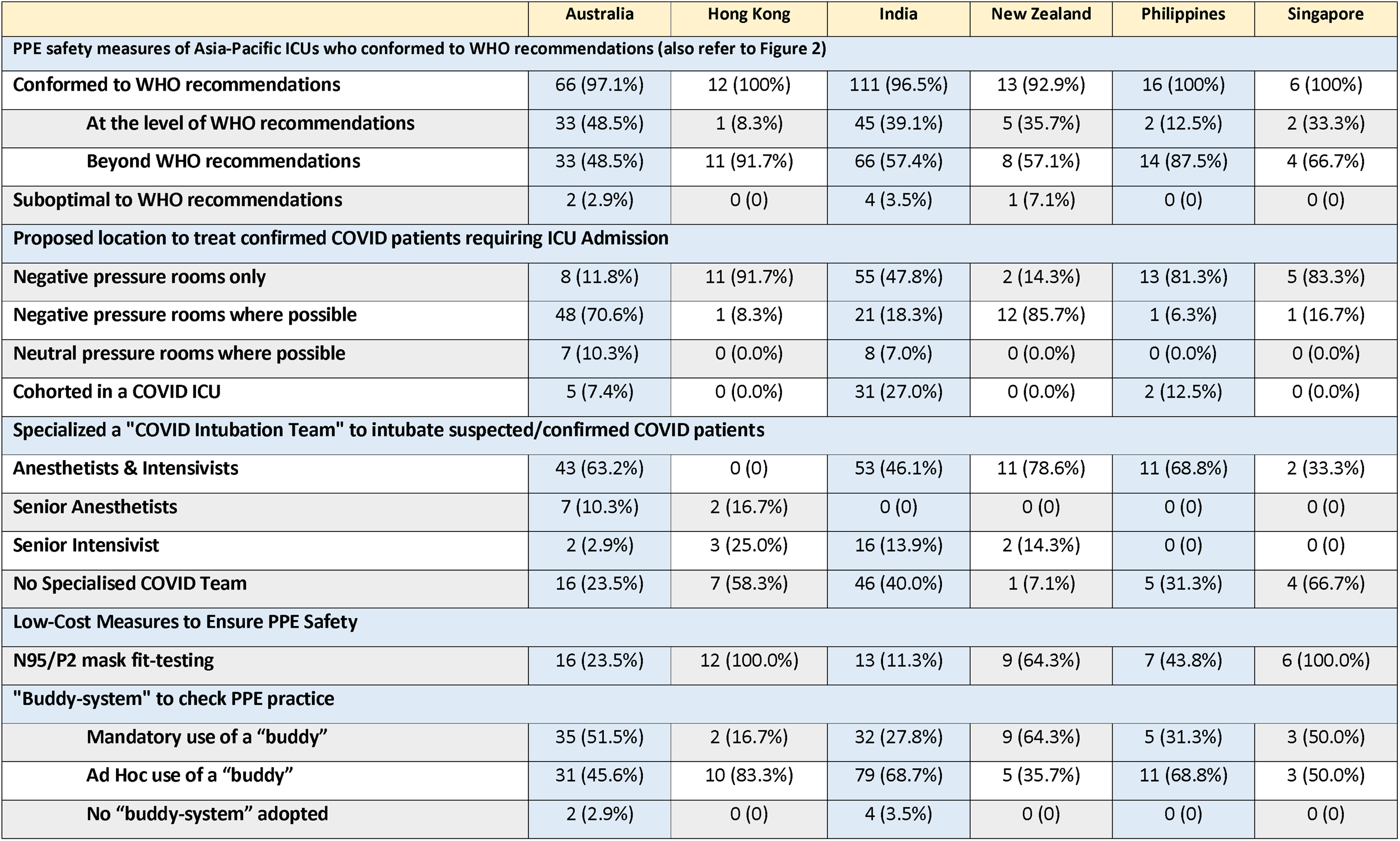
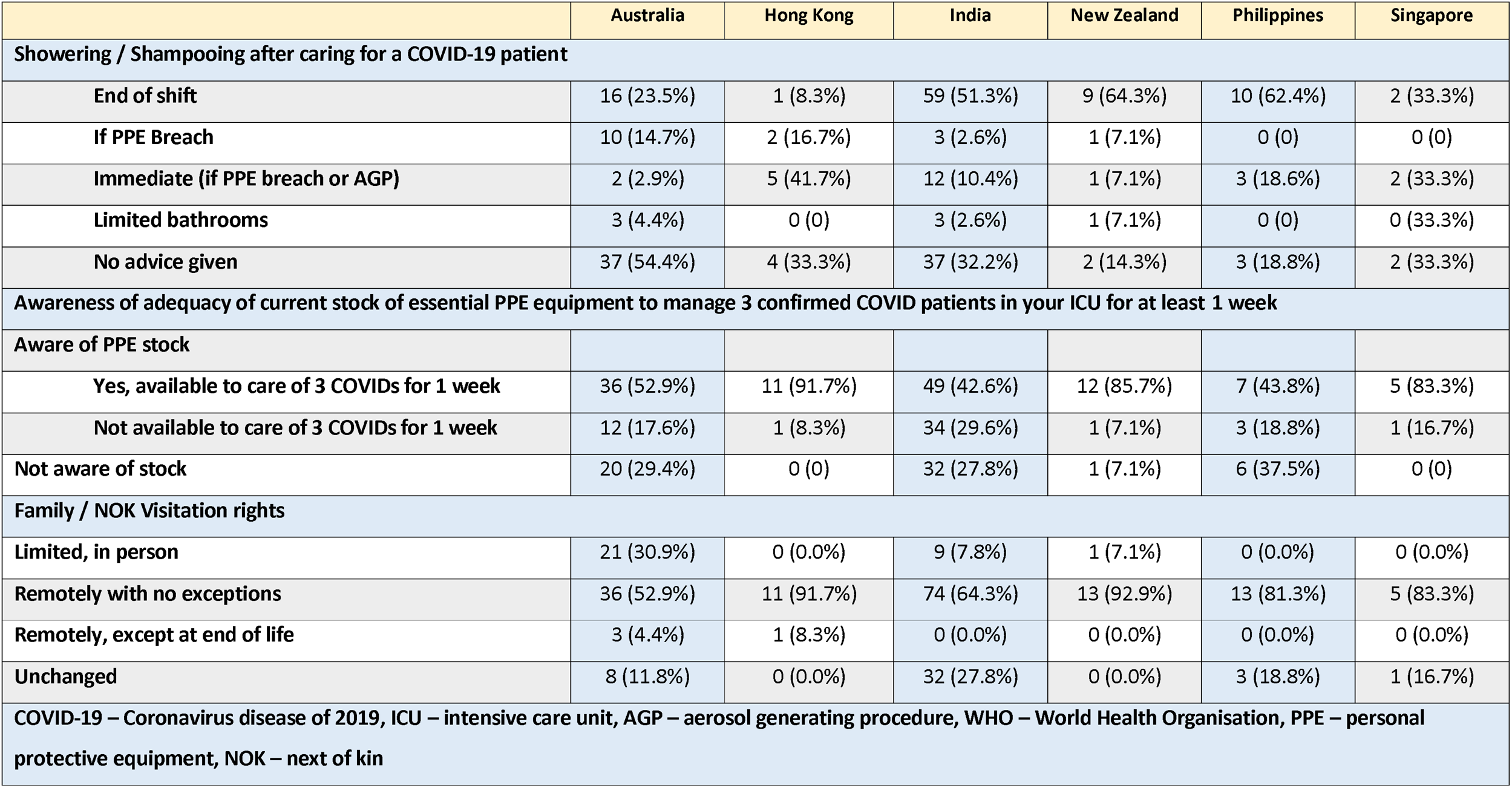
Key management strategies for Suspected/Confirmed COVID-19 patient

**Table 3.** Training processes for Activities Carrying High-Risk of Aerosol-Generation

|  | Daily or frequently | Not aware or not done | Occasional |
| --- | --- | --- | --- |
| **Intubating COVID-19 patients** |  |  |  |
| Australia | 40 (58.8) | 9 (13.2) | 19 (27.9) |
| Hong Kong | 4 (33.3) | 0 (0) | 8 (66.7) |
| India | 21 (18.3) | 52 (45.2) | 42 (36.5) |
| New Zealand | 11 (78.6) | 0 (0) | 3 (21.4) |
| Philippines | 4 (25.0) | 6 (37.5) | 6 (37.5) |
| Singapore | 3 (50.0) | 0 (0) | 3 (50.0) |
| **Transporting COVID-19 patients** |  |  |  |
| Australia | 15 (22.1) | 26 (38.2) | 27 (39.7) |
| Hong Kong | 1 (8.3) | 10 (83.3) | 1 (8.3) |
| India | 19 (16.5) | 60 (52.2) | 36 (31.3) |
| New Zealand | 7 (50.0) | 2 (14.3) | 5 (35.7) |
| Philippines | 4 (25.0) | 5 (31.3) | 7 (43.8) |
| Singapore | 1 (16.7) | 1 (16.7) | 4 (66.7) |
| **PPE donning and doffing** |  |  |  |
| Australia | 54 (79.4) | 1 (1.5) | 13 (19.1) |
| Hong Kong | 9 (75.0) | 0 (0) | 3 (25.0) |
| India | 48 (41.7) | 22 (19.1) | 45 (39.1) |
| New Zealand | 14 (100.0) | 0 (0) | 0 (0) |
| Philippines | 10 (62.5) | 1 (6.3) | 5 (31.3) |
| Singapore | 4 (66.7) | 0 (0) | 2 (33.3) |
| **PPE waste disposal for cleaners** |  |  |  |
| Australia | 25 (36.8) | 23 (33.8) | 20 (29.4) |
| Hong Kong | 4 (33.3) | 5 (41.7) | 3 (25.0) |
| India | 43 (37.4) | 36 (31.3) | 36 (31.3) |
| New Zealand | 6 (42.9) | 5 (35.7) | 3 (21.4) |
| Philippines | 9 (56.3) | 3 (18.8) | 4 (25.0) |
| Singapore | 2 (33.3) | 0 (0) | 4 (66.7) |
| **COVID-19 – coronavirus disease 2019** |  |  |  |

Regular training in donning/doffing was provided in all NZ ICUs, 79% in Australia, 75% in HK, 67% in Singapore, 62% in Philippines and 42% in India. Regular training for intra-hospital transport of COVID-19 patients ranged between 8% (HK) to 50% (NZ). Regular training on waste disposal for ICU cleaners ranged between 33% (HK/Singapore) and 56% (Philippines).

### PPE Practice (i.e., choice of equipment) - Table 2, Figure 2 and sFigure 1

224/231 (97%) either conformed to/exceeded the WHO recommendations. 3% did not report wearing masks. 38% conformed to the WHO recommendations of using medical masks for non-AGPs (“droplet precautions”) and limiting N95/P2 masks (i.e., airborne precautions) to AGPs alone. 59% used airborne precautions routinely, irrespective of AGP risk (range 48% in Australia to 92% in HK) (Figure 2).

**Figure 2.**
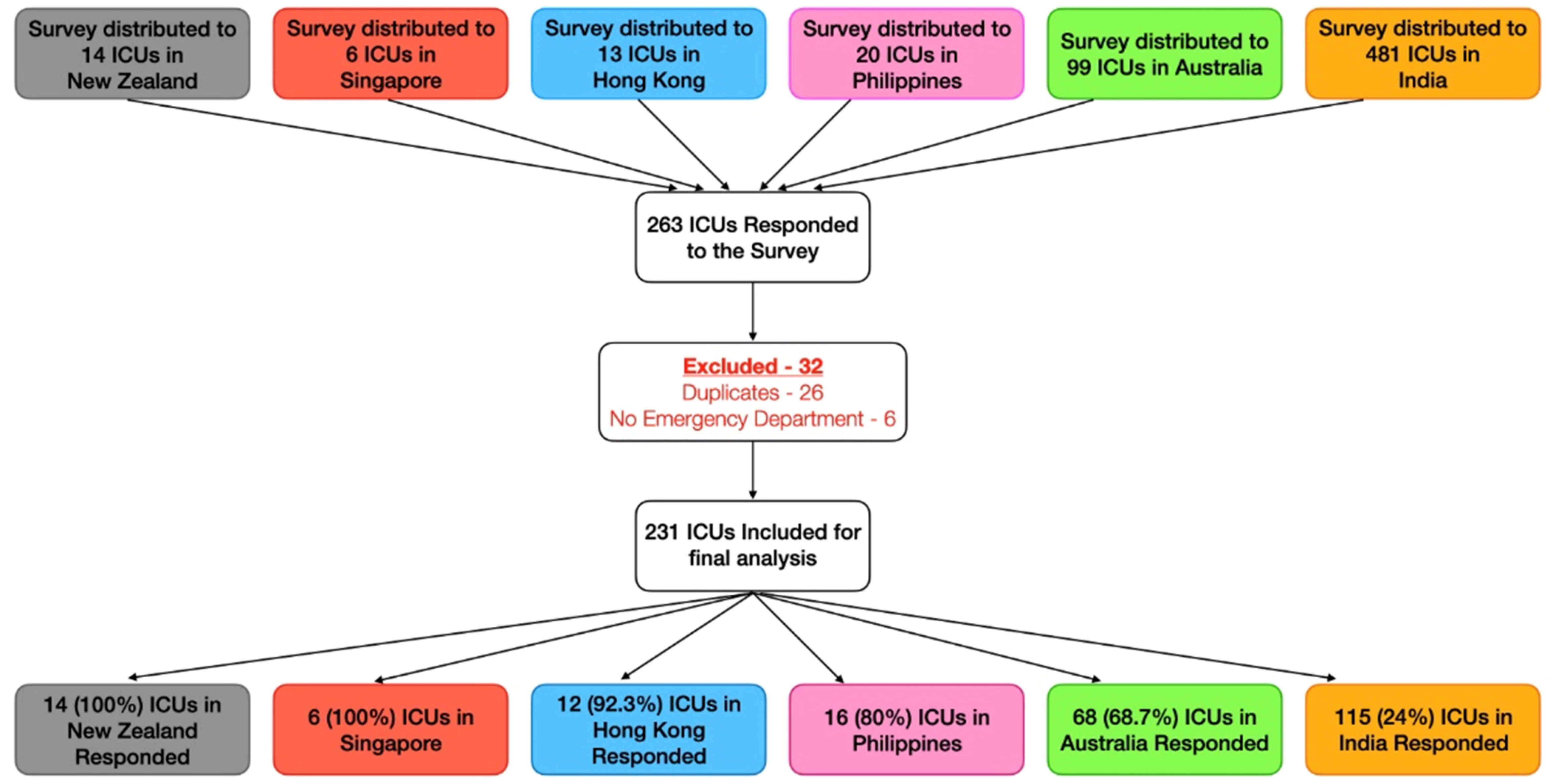
Figure 2a shows respiratory PPE practices reported by intensivists in the different countries. Figure 2b shows how this reported use conformed to the WHO guidelines. Key: PAPR - personal air-purifying respirators N95- N9 mask WHO-World Health Organisation

Only 6% used personal-air-purifying-respirators (PAPR), except in Singapore where half the ICUs used PAPR for AGPs. 35% ICUs used full body suits, with variations between countries – not used in NZ to 94% in Philippines.

sFigure 1 summarizes the use of head-covers/caps (71%), shoe-covers (45%), neck- covers (37%), hospital scrubs (58%) and impervious gowns (58%). Showering/shampooing hair were routine practice in 60% ICUs, typically after the shift (46%), and/or after PPE breach (15%) (Table 1).

27% ICUs performed N95/P2-mask fit-testing with quantitative/qualitative methods. There was wide inter-country variability, from 100% (HK, Singapore), 64% (NZ) to 11%-44% in the others.

Observers to monitor/checking on colleagues while donning/doffing PPE (“buddy- system”)[32] was mandatory practice in 64% in NZ ICUs, 51% in Australia and 50% in Singapore, and less common in other countries, ranging between 16%-31%.

### Disposition of COVID-19 patients in the ICU and modes of oxygen-therapy for non-intubated patients

There were variations on where the different ICUs countries managed COVID-19 patients (Table 3). Respondents from 37% ICUs (especially in HK and Singapore) stated using negative-pressure rooms exclusively. Overall, 58% were prepared to use non-negative-pressure rooms if necessary (i.e. neutral-pressure single rooms, dedicated/cohorted COVID-19 area). Only 1 respondent each from HK and Singapore reported using non-negative-pressure rooms.

Low-flow oxygen-therapy, high-flow nasal oxygenation (HFNO) and non-invasive ventilation (NIV) were reported as “not an option in COVID-19 patients” by 14%, 26% and 45% respondents respectively, particularly in Singapore and HK, where 80% avoided NIV, and 50% and 67% avoided HFNO respectively (Figure 3). Other respondents were prepared to use low-flow oxygen (39%), HFNO (45%) and NIV (34%) for patients in negative-pressure rooms or dedicated/cohorted areas.

**Figure 3.**
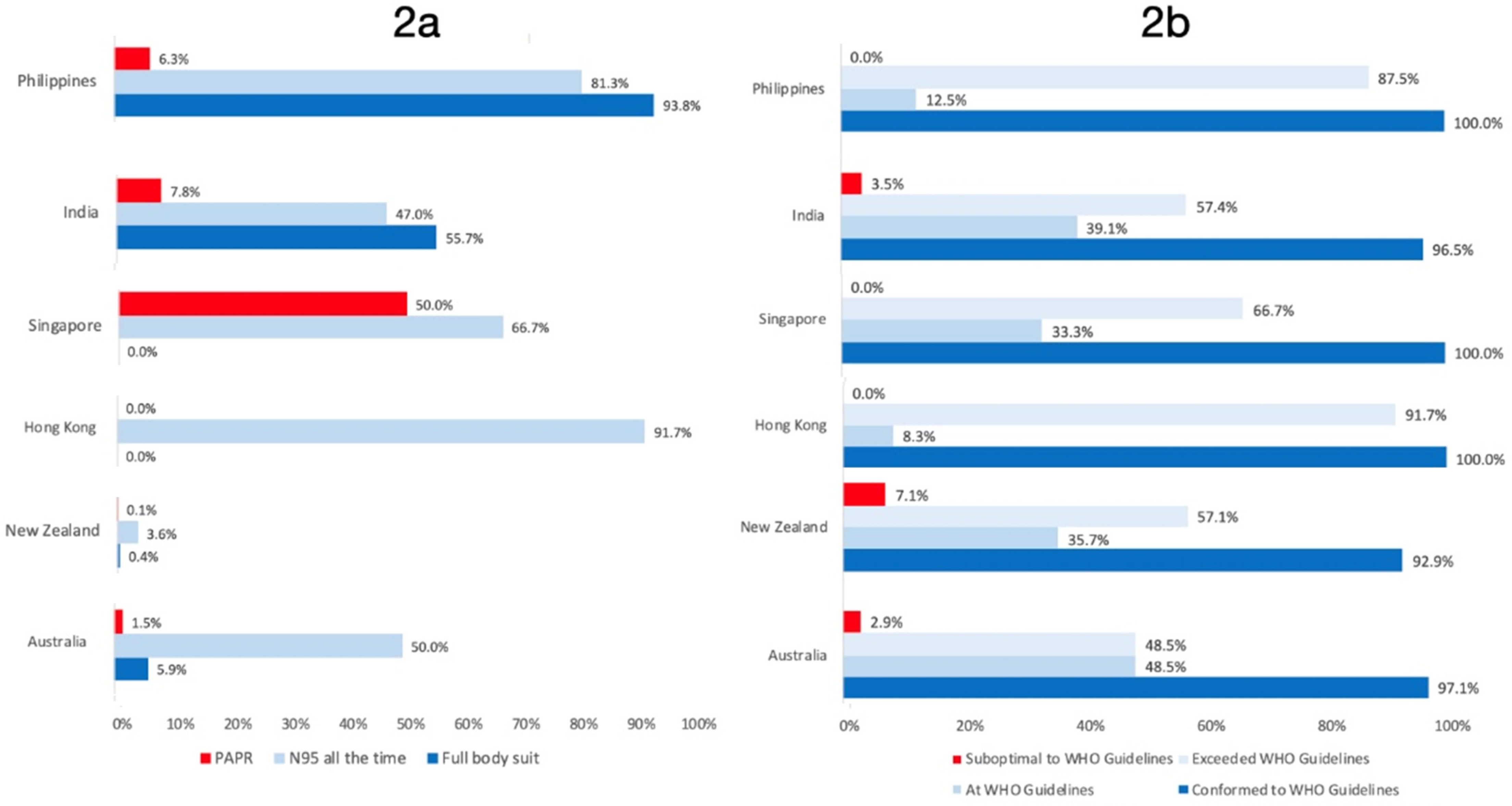
Disposition and management of a non-intubated COVID-19 patient Key: COVID-19 - coronavirus disease 2019

### Other aspects

Awareness of institutional PPE stocks being adequate to manage three COVID-19 patients for 1 week ranged from 41% (Australia) to 92% (HK) (Table 2).

Overall, 70% of respondents curbed family visits to the ICU (Table 2). Family communications were either by phone or videoconferencing. NZ, HK and Singapore ICUs all had reduced visiting rights, compared with 12-28% in other countries where policies were unchanged.

Overall, 28% of respondents reported feeling safe, many of them in NZ, Singapore and HK. 29% felt unsafe, and the remainder were neutral. Between 64%-83% of Singaporean, HK and NZ intensivists expressed confidence in their PPE-preparedness, while most intensivists in Australia, India and Philippines felt that their PPE-preparedness were suboptimal (sTable

## Discussion

### Statement of principal findings

To our knowledge, this multinational survey is the first to systematically evaluate ICU PPE- preparedness for the COVID-19 pandemic. Although the overwhelming majority of ICUs conformed to the WHO recommendations for PPE-usage, there were marked variations in the level of PPE-preparedness. ICUs in NZ, HK and Singapore appeared to have the most well-developed systems, training and/or practice compared to ICUs across Australia, India and Philippines, which had inconsistent systems. There were variations between countries in every aspect of PPE-preparedness namely, PPE-training, PPE stock-awareness, negative-pressure room use, use of HFNO/NIV for non-intubated patients, “buddy-systems”, team-training exercises and showering practices.

### Variations in PPE-training

The most important and immediately remediable concern identified was the suboptimal training for ICU-HCWs, which may lead to suboptimal PPE-practice. Although regular training on PPE donning and doffing was offered in most ICUs, training for other aerosol-generating activities was inconsistent in all countries, with NZ being better than the others. Such ICUs may aim to emulate the PPE-training reported by NZ respondents, and immediately initiate training sessions on N95/P2 mask fit-testing, donning/doffing PPE, AGP-minimisation strategies, specialised intubation teams, transport, cleaning/waste disposal, and low-cost but effective mechanisms such as buddy-systems. These will improve safety and team-bonding.[8,26,32]

### Disposition of COVID-19 patients in the ICU and Modes of Oxygen-Therapy for Non-Intubated Patients

Compared to other countries, more intensivists from HK and Singapore reported that they would exclusively use negative-pressure rooms to manage COVID-19 patients. This may reflect the increased availability of such rooms in their ICUs, in turn demonstrating better pandemic-preparedness following the SARS-CoV-1 pandemic. Despite this, respondents from these 2 countries commonly avoided using HFNO and NIV, while intensivists in other countries used them more commonly. The use of HFNO and NIV in negative-pressure rooms or cohorted areas is recommended by several PPE guidelines, provided HCWs employ PPE with airborne-precautions.[15,33] Inappropriate avoidance of these therapies may result in unnecessary intubation or palliation of patients, hence needs reconsideration.

### Variations in PPE-practice across countries; Potential effects of conflicting PPE-recommendations

The variations in ICU PPE-practice across the countries surveyed may partly be due to the startling multitude of international, national, regional, local/institutional and even departmental PPE guidelines, sometimes making contradictory recommendations (sTable 1). For instance, when managing COVID-19 patients, one-third of respondents followed the WHO “Rational Use of PPE” recommendations[5] of reserving airborne precautions (i.e., N95/P2 masks) exclusively for AGPs, while 60% were in line with the ANZICS recommendations of routinely using N95/P2 masks, irrespective of AGPs. There were variations between and within countries, with airborne precautions being more common in HK and Singapore. There is pre-COVID-19 evidence that routine airborne precautions may be more protective than targeted airborne-precautions.[19,20,34] However, the optimal strategy to strike a balance between conserving N95/P2 masks and ensuring ICU-HCW safety is unclear. Regardless, since conflicting PPE-recommendations have the potential to cause confusion and errors, [35] health administrators in a region/state/country must ensure a clear and consistent guideline across their jurisdiction to promote consistency in training/practice and build staff confidence. Health advisory organisations may attempt to unify their PPE recommendations to minimise practice-variation.

### Other aspects

It is concerning that many intensivists were unaware of their PPE stock adequacy. This demands urgent attention. Administrators may consider implementing innovative solutions to keep track of each hospital’s current and future PPE stock, as has been done in Australia/NZ.[36]

Our survey found that very few respondents reported using neck and shoe covers. Also, most PPE-guidelines do not incorporate these in their recommendations. As the soles of HCWs’ shoes might function as carriers in spreading the virus, ICU-HCWs may consider stricter protective measures (e.g., caps, shoe and neck covers).[37]

Regarding family visitation, almost one-third of ICUs reported unchanged practices, which may potentially deplete PPE stocks, but also expose family members to infection.[37] As employed by NZ, HK and Singapore ICUs, phone or video-conferencing communication solutions may be suitable alternatives.

Finally, it was clear that good PPE-preparedness resulted in better perceptions of ICU-HCW safety. More intensivists from ICUs with homogeneous PPE-practices (HK, Singapore and NZ) felt safer, compared to intensivists from the other countries.

### Strengths and weaknesses

The strengths of this survey included a robust process to develop the questionnaire. Responses were limited to one intensivist/ICU; non-medical respondents were excluded to keep the respondents homogeneous. Both well-resourced and less-resourced countries were included. Even in the midst of the COVID-19 pandemic, the response rate in all countries was high, except India, where responses were obtained widely from 23/27 states.

Our study has several limitations. Inherent to any survey, the submissions were self-declared statements without independent corroboration. Given the rapid evolution of PPE-preparedness, it is possible that the issues identified during the survey have already been addressed. The random selection of participants, coupled with the exclusion of non-medical HCWs, may induce reporting bias. The survey did not evaluate other AGPs like prone positioning, cardiac arrest, tracheostomy and bronchoscopies, partly due to unresolved ethical dilemmas (cardiac arrest) and partly to ensure feasibility of completion. The choice of the broad and pragmatic WHO guideline over ICU-specific guidelines (ANZICS) as the reference standard for ICU PPE practice is questionable but was done since it is more applicable to a multinational setting and does have a section on PPE for AGPs. Finally, despite the wide geographical spread of respondents, the low response rate in India and the small number of Philippines ICUs may limit the applicability of the results in those countries.

### Unanswered questions and future research

Although the link between PPE-preparedness and ICU-HCW infections is plausible, the association/causal link may be better evaluated using case-control or retrospective cohort studies to compare HCW-infection rates between countries with variable training/practices. A follow-up survey may help evaluate any changes in PPE-practice/training in the ICUs that were surveyed.

### Meaning of the study

The survey identified several deficiencies in PPE-preparedness across Asia-Pacific ICUs that demand the urgent attention of administrators and policymakers to institute corrective steps to ensure HCW safety.

## Conclusions

The survey found that most intensivists from six Asia-Pacific countries showed good awareness of the WHO PPE-guidelines by either conforming to/exceeding the recommendations. Despite this, there were widespread variabilities across ICUs and countries in several domains, particularly related to PPE-training and preparedness. Standardising PPE guidelines may translate to better training, better compliance and policies that improve HCW safety. Adopting low-cost approaches such as buddy-systems should be encouraged. More importantly, better pandemic preparedness and building systems with deeply embedded culture of safety is essential to ensure the safety and well-being of HCWs during such pandemics.

## Data Availability

Data available on request by email

## Acknowledgements

The authors thank Dr Adam Howard, Intensivist, Royal Perth Hospital, Western Australia, Dr Ross Freebairn, Intensivist, Hawkes Bay Hospital, New Zealand and Dr Paul Young, Wellington Hospital, New Zealand for their valuable inputs. Dr Shekar acknowledges research support from Metro North Hospital and Health Service. Dr Haji would like to acknowledge Dr. Prashant Kumar Editor ‘Critical Care WA articles’ HOD Critical Care, KHNI, Delhi - NCR for his help with the distribution of the survey in India.

## Author Contributions

Listed in Title Page

## Copyright/license for publication

The Corresponding Author has the right to grant on behalf of all authors and does grant on behalf of all authors, a worldwide license to the Publishers and its licensees in perpetuity, in all forms, formats and media (whether known now or created in the future), to i) publish, reproduce, distribute, display and store the Contribution, ii) translate the Contribution into other languages, create adaptations, reprints, include within collections and create summaries, extracts and/or, abstracts of the Contribution, iii) create any other derivative work(s) based on the Contribution, iv) to exploit all subsidiary rights in the Contribution, v) the inclusion of electronic links from the Contribution to third party material where—ever it may be located; and, vi) license any third party to do any or all of the above.

## Transparency Declaration

The lead author and the manuscript’s guarantor, Dr Arvind Rajamani, affirms that the manuscript is an honest, accurate, and transparent account of the study being reported; that no important aspects of the study have been omitted; and that any discrepancies from the study as originally planned (and, if relevant, registered) have been explained.

## Funding source – Nil

### Patient and Public Involvement

Patients or the public WERE NOT involved in the design, or conduct, or reporting, or dissemination plans of our research

